## Supplementary material for "Risk assessment for airborne disease transmission by poly-pathogen aerosols": S1_appendix

### S1 Appendix. Model Solution Derivation

#### General Solution

The model is a finite coupled system of linear inhomogeneous ODEs for any fixed $d_0$. In matrix-vector form, it is

$$\frac{d\vec{n}}{dt} = \mathbf{A}(d_0,t)\vec{n}(d_0,t) + \vec{\beta}(d_0,t) \quad , \tag{1}$$

where $\vec{n}(d_0,t)$ and $\vec{\beta}(t)$ are the $n_k(d_0 t)$ and $\beta_k(d_0,t)$ for $k > 0$ in vector form and

$$\mathbf{A} \equiv \begin{bmatrix} -\alpha(d_0,t) - \gamma(t) & 2\gamma(t) & & & \\ & -\alpha(d_0,t) - 2\gamma(t) & 3\gamma(t) & & \\ & & \ddots & \ddots & \\ & & & \ddots & M_c\gamma(t) \\ & & & & -\alpha(d_0,t) - M_c\gamma(t) \end{bmatrix} \quad . \tag{2}$$

is an upper bidiagonal $M_c \times M_c$ square matrix.

It is well-known that the complementary solution (the one for when $\vec{\beta} = \vec{0}$) is $\vec{n}_C(d_0,t) = \mathbf{X}(d_0,d_0,t)\mathbf{X}^{-1}(d_0,t_0)\vec{n}_0$ where $\mathbf{X}(d_0,t) = \exp\left[\int^t \mathbf{A}(d_0,x)dx\right]$ if $\mathbf{A}(d_0,t)$ commutes with itself at all combinations of time at fixed $d_0$ ($\mathbf{A}(d_0,x)\mathbf{A}(d_0,y) = \mathbf{A}(d_0,y)\mathbf{A}(d_0,x) \;\; \forall \;\; x,y \in \mathbb{R}$) [1]. As we will later show from its diagonalization, $\mathbf{A}$ commutes with itself for any two combinations of time with fixed $d_0$. Let its diagonalization be $\mathbf{A} = \mathbf{B}\mathbf{\Lambda}\mathbf{B}^{-1}$, which is gotten via eigenvalue decomposition. We will use the notation $\mathbf{c}_{m,k}$ to denote the $m$'th row and $k$'th column of a matrix $\mathbf{C}$.

It can be shown that the eigen values of a bidiagonal matrix are its diagonal elements. The eigenvalue matrix $\mathbf{\Lambda}$ is a diagonal matrix where the $k$'th diagonal element is the $k$'th eigen value. Putting them in the same order as the diagonal elements of $\mathbf{A}$, they are

$$\ell_k(d_0,t) = -\alpha(d_0,t) - k\gamma(t) \quad . \tag{3}$$

Except in the degenerate but trivial case $\gamma(t) = 0$, there are $M_c$ distinct eigenvalues (not degenerate), and in the degenerate case $\mathbf{A} = -\alpha\mathbf{I}$ and is trivially diagonalized ($\mathbf{\Lambda} = \mathbf{A}$ and $\mathbf{B} = \mathbf{B}^{-1} = \mathbf{I}$). So $\mathbf{A}$ has $M_c$ linearly independent eigen vectors and is therefore diagonalizable. To complete the diagonalization, we need to find the eigen vector $\vec{b}_k(d_0, t)$ for each eigen value by solving

$$\mathbf{A}\vec{b}_k = \ell_k \vec{b}_k \quad . \tag{4}$$

We can begin construction of the eigenvector by setting all elements $m > k$ to zero and element $m = k$ to one. For elements $m \geq k$, the eigen value-vector relationship above trivially holds. For element $m < k$, this becomes the following recursive relationship for the $m$'th row in terms of row $m + 1$

$$\begin{aligned} \ell_m b_{m,k} + (m+1)\gamma b_{m+1,k} &= \ell_k b_{m,k} \\ b_{m,k} &= -\left(\frac{m+1}{k-m}\right) b_{m+1,k} \quad , \end{aligned} \tag{5}$$

which starts with $b_{k,k} = 1$. From this, element $m < k$ is

$$b_{m,k} = (-1)^{k-m} \frac{(m+1)(m+2)\cdots k}{(k-m)(k-m-1)\cdots 1} = (-1)^{k-m} \binom{k}{m} \quad , \tag{6}$$

where $\binom{k}{m} = k!/(m!(k-m)!)$ is the notation for the binomial coefficient $k$ choose $m$. Conveniently, this means that all the elements of the eigen vectors are integers and there is no dependence on $t$ nor $d_0$. The full eigen vector matrix is

$$b_{m,k} = \begin{cases} (-1)^{k-m}\binom{k}{m} & \text{if } m < k \quad , \\ 1 & \text{if } m = k \quad , \\ 0 & \text{if } m > k \quad , \end{cases} \tag{7}$$

where the first case is technically also true along the diagonal ($m = k$) since it is equal to one but it is more clear to write the diagonal elements explicitly as one. The inverse of $\mathbf{B}$ is equal to the matrix $\mathbf{B}$ but with all elements replaced by their absolute values. The elements of $\mathbf{B}^{-1}$ are thus

$$\left(b^{-1}\right)_{m,k} = \begin{cases} \binom{k}{m} & \text{if } m < k \quad , \\ 1 & \text{if } m = k \quad , \\ 0 & \text{if } m > k \quad . \end{cases} \tag{8}$$

This was originally guessed after looking at the numerically computed inverse, but we will prove it here. Consider the $m$'th row and the $k$'th column of $\mathbf{B}\mathbf{B}^{-1}$ which is just $\left(\mathbf{B}\mathbf{B}^{-1}\right)_{m,k} = \sum_{p=1}^{M_c} b_{m,p}\left(b^{-1}\right)_{p,k}$. The elements of the sum are zero unless $m \leq p \leq k$, which means that $\left(\mathbf{B}\mathbf{B}^{-1}\right)_{m,k} = 0$ for $m > k$. For $m \leq k$, the zero elements at the beginning and end of the sum drop and we get a sum between $m$ and $k$ that is

$$\begin{aligned}\left(\mathbf{B}\mathbf{B}^{-1}\right)_{m,k} &= \sum_{p=m}^{k}(-1)^{p-m}\binom{p}{m}\binom{k}{p}\\ &= \sum_{q=0}^{s}(-1)^{p}\binom{q+m}{m}\binom{k}{q+m}\\ &= \sum_{q=0}^{s}(-1)^{q}\frac{k!}{m!q!(k-q-m)!}\\ &= \frac{k!}{m!s!}\sum_{q=0}^{s}(-1)^{q}\binom{s}{q}\\ &= \begin{cases}1 & \text{if } m=k \quad ,\\ 0 & \text{if } m<k \quad ,\end{cases}\end{aligned} \tag{9}$$

where we have defined $q \equiv p-m$ and $s \equiv k-m$, the last sum is trivially equal to 1 if $m=k$ since then $s=0$ and only the first term is present, and the sum is zero for $s>0$ ($k>m$) as proven by Aupetit (Eq 7 in Appendix) [2]. All non-diagonal elements are zero and all diagonal elements are one, meaning that we have the identity matrix $\mathbf{I}$. Thus Eq 8 is indeed the matrix inverse of Eq 7.

Thus we have diagonalized $\mathbf{A}$. We can see that while the eigen values depend on $t$ and $d_0$ if $\alpha$ or $\gamma$ do; $\mathbf{B}$ (and therefore $\mathbf{B}^{-1}$) do not and are therefore constant. Note that $\mathbf{A}(d_0,t)$, $\mathbf{B}$, $\mathbf{B}^{-1}$, and $\mathbf{\Lambda}(d_0,t)$ are all upper-triangular matrices. Now we can show that $\mathbf{A}$ commutes with itself for any two times but fixed $d_0$. For two arbitrary times $x, y \in \mathbb{R}$, the product is

$$\begin{aligned}\mathbf{A}(x)\mathbf{A}(y) &= \mathbf{B}\mathbf{\Lambda}(x)\mathbf{B}^{-1}\mathbf{B}\mathbf{\Lambda}(y)\mathbf{B}^{-1}\\ &= \mathbf{B}\mathbf{\Lambda}(y)\mathbf{\Lambda}(x)\mathbf{B}^{-1}\\ &= \mathbf{B}\mathbf{\Lambda}(y)\mathbf{B}^{-1}\mathbf{B}\mathbf{\Lambda}(x)\mathbf{B}^{-1}\\ &= \mathbf{A}(y)\mathbf{A}(x) \quad ,\end{aligned}$$

since all diagonal matrices, such as $\mathbf{\Lambda}(t)$, commute with each other. Thus $\mathbf{A}$ commutes with itself for any two times (but fixed $d_0$). Since $\mathbf{A}$ commutes with itself for any two times, $\int^t \mathbf{A}(d_0,x)dx$ also commutes itself for any two times (integral can be turned into an infinite Riemann sum over $\mathbf{A}$ which shows that it commutes with itself). Then the multiplied matrix exponentials $\exp\left[\int^t \mathbf{A}(d_0,x)dx\right]\exp\left[-\int^t \mathbf{A}(d_0,x)dx\right]$ become the matrix exponential of their sums. Choosing the constant of integration such that $\vec{n}_C(d_0,0)=\vec{n}_0(d_0)$, we get for the complete complementary solution

$$\vec{n}_C(d_0,t) = \exp\left[\int_{t_0}^{t}\mathbf{A}(d_0,x)dx\right]\vec{n}_0 \quad . \tag{10}$$

Then the general solution to Eq (1) is

$$\vec{n}(d_0,t) = \exp\left[\int_{t_0}^{t}A(d_0,x)dx\right]\vec{n}_0(d_0) + \int_{t_0}^{t}\exp\left[\int_{s}^{t}A(d_0,x)dx\right]\vec{\beta}(d_0,s)ds \quad , \tag{11}$$

which we got by guessing based on the equivalent solution for a single equation rather than a system and then checking it (checking it is straightforward).

Now we need the matrix exponential $\exp\left[\int_s^t \mathbf{A}dx\right]$ for some scalar $s$. All $t$ and $d_0$ dependence in the diagonalization of $\mathbf{A}$ is confined to the eigen value matrix $\mathbf{\Lambda}(d_0, t)$, which greatly simplifies finding this matrix exponential. First, we can rewrite the eigenvalue matrix as

$$\mathbf{\Lambda}(d_0, t) = -\alpha(d_0, t)\mathbf{I} - \gamma(t)\mathbf{G} \quad , \tag{12}$$

where $\mathbf{G}$ is the diagonal matrix

$$\mathbf{G} = \begin{bmatrix} 1 & & \\ & \ddots & \\ & & M_c \end{bmatrix} \quad . \tag{13}$$

Since $\mathbf{A}$ is diagonalized, all diagonal matrices commute with each other with respect to multiplication, $\mathbf{B}$ is not a function of time, and the matrix exponential of the sum of two matrices is the product of their exponentials if they commute; the matrix exponential of $\int_s^t \mathrm{A}(d_0, s)dx$ is

$$\begin{aligned} \exp\left[\int_s^t \mathrm{A}(d_0, x)dx\right] &= \mathbf{B}\exp\left[\int_s^t \mathbf{\Lambda}(d_0, x)dx\right]\mathbf{B}^{-1} \\ &= \mathbf{B}\exp\left[-\mathbf{I}\int_s^t \alpha(x)dx - \mathbf{G}\int_s^t \gamma(x)dx\right]\mathbf{B}^{-1} \\ &= \mathbf{I}\exp\left[-\int_s^t \alpha(d_0, x)dx\right]\mathbf{B}\exp\left[-\mathbf{G}\int_s^t \gamma(x)dx\right]\mathbf{B}^{-1} \\ &= u(d_0, t, s)\mathbf{H}(t, s) \quad , \end{aligned} \tag{14}$$

where

$$u(d_0, t, s) = \exp\left[-\int_s^t \alpha(d_0, x)dx\right] \quad , \tag{15}$$

$$H(t, s) = \mathbf{B}\exp\left[-\mathbf{G}\int_s^t \gamma(x)dx\right]\mathbf{B}^{-1} \quad . \tag{16}$$

We can put this into the general solution for the system of ODEs from Eq (11) to get

$$\vec{n}(d_0, t) = u(d_0, t, t_0)\mathbf{H}(t, t_0)\vec{n}_0(d_0) + \int_{t_0}^t u(d_0, t, s)\mathbf{H}(t, s)\vec{\beta}(d_0, s)ds \quad . \tag{17}$$

Now we must determine $\mathbf{H}$. Since $\mathbf{G}$ is a diagonal matrix

$$\exp\left[-\mathbf{G}\int_s^t \gamma(x)dx\right] = \begin{bmatrix} e^{-\int_s^t \gamma(x)dx} & & \\ & \ddots & \\ & & e^{-M_c\int_s^t \gamma(x)dx} \end{bmatrix} \quad . \tag{18}$$

When this multiplies with $\mathbf{B}$, it essentially scales the columns by the diagonal elements. The resulting matrix is an upper triangular matrix. For the $m$'th row and $k$'th column, $h_{m,k}(t, s) = 0$ if $m > k$; and for $m \leq k$ it is

$$
\begin{aligned}
h_{m,k}(t,s) &= \sum_{i=m}^{k} b_{m,i}(b_{i,k})^{-1} \exp\left[-i\int_s^t \gamma(x)dx\right] \\
&= \sum_{i=m}^{k} \frac{(-1)^{i-m}k!}{m!(i-m)!(k-i)!} \exp\left[-i\int_s^t \gamma(x)dx\right] \\
&= \sum_{p=0}^{k-m} \frac{(-1)^{p}k!}{m!p!(k-m-p)!} \exp\left[-(p+m)\int_s^t \gamma(x)dx\right] \\
&= \frac{k!}{m!(k-m)!} \sum_{p=0}^{k-m} (-1)^p \binom{k-m}{p} \exp\left[-(p+m)\int_s^t \gamma(x)dx\right] \\
&= \binom{k}{m} \sum_{p=0}^{k-m} (-1)^p \binom{k-m}{p} \exp\left[-(p+m)\int_s^t \gamma(x)dx\right] \quad , \qquad (19) \\
&= \binom{k}{m} \exp\left[-m\int_s^t \gamma(x)dx\right] \sum_{p=0}^{k-m} \binom{k-m}{p} \left[-\exp\left[-\int_s^t \gamma(x)dx\right]\right]^p \\
&= \binom{k}{m} \exp\left[-m\int_s^t \gamma(x)dx\right] \left[1-\exp\left[-\int_s^t \gamma(x)dx\right]\right]^{k-m} \quad , \qquad (20)
\end{aligned}
$$

where the last step uses the definition of binomial coefficients. One could use either Eq (19) or (20) to calculate the elements or do further derivations.

Dropping out of vector form, we can now write Eq (17) for each multiplicity as

$$
\begin{aligned}
n_k(d_0,t) = u(d_0,t,t_0) \exp\left[-k\int_{t_0}^t \gamma(x)dx\right] \\
\bullet \sum_{p=k}^{M_c} \binom{p}{k} n_{0,p}(d_0) \left[1-\exp\left[-\int_{t_0}^t \gamma(x)dx\right]\right]^{p-k} \\
+ \sum_{p=k}^{M_c} \binom{p}{k} \int_{t_0}^t u(d_0,t,s)\beta_p(d_0,s) \\
\bullet \exp\left[-k\int_s^t \gamma(x)dx\right] \left[1-\exp\left[-\int_s^t \gamma(x)dx\right]\right]^{p-k} ds \quad , \qquad (21)
\end{aligned}
$$

or with $u(d_0,t,t_0)$ substituted out as

$$
\begin{aligned}
n_k(d_0,t) = \exp\left[-\int_{t_0}^t \alpha(d_0,x)dx\right] \exp\left[-k\int_{t_0}^t \gamma(x)dx\right] \\
\bullet \sum_{p=k}^{M_c} \binom{p}{k} n_{0,p}(d_0) \left[1-\exp\left[-\int_{t_0}^t \gamma(x)dx\right]\right]^{p-k} \\
+ \sum_{p=k}^{M_c} \binom{p}{k} \int_{t_0}^t \beta_p(d_0,s) \\
\bullet \exp\left[-\int_s^t \alpha(d_0,x)dx\right] \exp\left[-k\int_s^t \gamma(x)dx\right] \left[1-\exp\left[-\int_s^t \gamma(x)dx\right]\right]^{p-k} ds \quad . \qquad (22)
\end{aligned}
$$

#### Solution for Coefficients Constant in Time

Eq (22) can be simplified if $\alpha$, $\vec{\beta}$, and/or $\gamma$ are constant with respect to time (or approximately so). If $\alpha(d_0)$ is constant with respect to time, then

$$u(d_0, t, s) = e^{-(t-s)\alpha(d_0)} \quad . \tag{23}$$

If $\gamma$ is constant (it has no $d_0$ dependence, so being constant with respect to time makes it a constant outright), then Eq (19) and (20) for expressing the upper triangle of $\mathbf{H}$ become

$$h_{m,k}(t,s) = \binom{k}{m} \sum_{p=0}^{k-m} (-1)^p \binom{k-m}{p} e^{-(p+m)(t-s)\gamma} \quad , \tag{24}$$

$$h_{m,k}(t,s) = \binom{k}{m} e^{-m(t-s)\gamma} \left[1 - e^{-(t-s)\gamma}\right]^{k-m} \quad . \tag{25}$$

If $\alpha$, $\vec{\beta}$, and $\gamma$ are all constant with respect to time; then for multiplicity $k$,

$$\begin{aligned}
&\left[\int_{t_0}^{t} u(d_0,t,s)\, \mathbf{H}(t,s)\vec{\beta}(d_0,s) ds\right]_k \\
&= \sum_{i=k}^{M_c} \binom{i}{k} \beta_i(d_0) \int_{t_0}^{t} e^{-(\alpha(d_0)+k\gamma)(t-s)} \left[1 - e^{-(t-s)\gamma}\right]^{i-k} ds \\
&= \sum_{i=k}^{M_c} \binom{i}{k} \beta_i(d_0) \sum_{p=0}^{i-k} \binom{i-k}{p} \int_{t_0}^{t} e^{-(\alpha(d_0)+k\gamma)(t-s)} \left[-e^{-(t-s)\gamma}\right]^{p} ds \\
&= \sum_{i=k}^{M_c} \binom{i}{k} \beta_i(d_0) \sum_{p=0}^{i-k} \binom{i-k}{p} (-1)^p \int_{t_0}^{t} e^{-[\alpha(d_0)+(k+p)\gamma](t-s)} ds \\
&= \sum_{i=k}^{M_c} \binom{i}{k} \beta_i(d_0) \sum_{p=0}^{i-k} \binom{i-k}{p} \frac{(-1)^p}{\alpha(d_0)+(k+p)\gamma} \\
&\quad \bullet \left[1 - e^{-[\alpha(d_0)+(k+p)\gamma](t-t_0)}\right] \quad .
\end{aligned} \tag{26}$$

Then if $\alpha$, $\vec{\beta}$, and $\gamma$ are all constant; the general solution from Eq (21) becomes

$$\begin{aligned}
n_k(d_0,t) &= e^{-(\alpha(d_0)+k\gamma)(t-t_0)} \sum_{i=k}^{M_c} \binom{i}{k} n_{0,i}(d_0) \left[1 - e^{-(t-t_0)\gamma}\right]^{i-k} \\
&+ \sum_{i=k}^{M_c} \binom{i}{k} \beta_i(d_0) \sum_{p=0}^{i-k} \binom{i-k}{p} \frac{(-1)^p}{\alpha(d_0)+(k+p)\gamma} \left[1 - e^{-[\alpha(d_0)+(k+p)\gamma](t-t_0)}\right] \quad .
\end{aligned} \tag{27}$$

The concentration density as $t \to \infty$ is

$$n_{\infty,k}(d_0) = \sum_{i=k}^{M_c} \binom{i}{k} \beta_i(d_0) \sum_{p=0}^{i-k} \binom{i-k}{p} \frac{(-1)^p}{\alpha(d_0)+(k+p)\gamma} \quad . \tag{28}$$

This term is present in Eq (27), so we can re-express it as

$$n_k(d_0,t) = n_{\infty,k}(d_0) + e^{-(\alpha(d_0)+k\gamma)(t-t_0)} \sum_{i=k}^{M_c} \binom{i}{k} \Bigg\{ n_{0,i}(d_0)\left[1 - e^{-(t-t_0)\gamma}\right]^{i-k} - \beta_i(d_0) \sum_{p=0}^{i-k} \binom{i-k}{p} \frac{(-1)^p e^{-p(t-t_0)\gamma}}{\alpha(d_0)+(k+p)\gamma} \Bigg\} \quad , \tag{29}$$

Calculation of the average aerosol dose requires the time integral of $n_k$. Completing the integral on all but the last term (will do an alternative evaluation of it later), it is

$$\begin{aligned} \int_{t_0}^{t} n_k(d_0,v)dv &= (t-t_0)n_{\infty,k}(d_0) \\ &+ \sum_{i=k}^{M_c} \binom{i}{k} n_{0,i}(d_0) \sum_{p=0}^{i-k} \binom{i-k}{p} \frac{(-1)^p}{\alpha(d_0)+(k+p)\gamma} \left[1 - e^{-[\alpha(d_0)+(k+p)\gamma](t-t_0)}\right] \\ &- \int_{t_0}^{t} dv\, e^{-(\alpha(d_0)+k\gamma)(v-t_0)} \sum_{i=k}^{M_c} \binom{i}{k} \beta_i(d_0) \sum_{p=0}^{i-k} \binom{i-k}{p} \frac{(-1)^p e^{-p(v-t_0)\gamma}}{\alpha(d_0)+(k+p)\gamma} \quad . \end{aligned} \tag{30}$$

And if we completely evaluate the last integral,

$$\begin{aligned} \int_{t_0}^{t} n_k(d_0,v)dv &= (t-t_0)n_{\infty,k}(d_0) \\ &+ \sum_{i=k}^{M_c} \binom{i}{k} \sum_{p=0}^{i-k} \binom{i-k}{p} \frac{(-1)^p}{\alpha(d_0)+(k+p)\gamma} \Bigg\{ n_{0,i}(d_0)\left[1 - e^{-[\alpha(d_0)+(k+p)\gamma](t-t_0)}\right] \\ &+ \beta_i(d_0) \left( \frac{e^{-[\alpha(d_0)+(k+p)\gamma](t-t_0)} - 1}{\alpha(d_0)+(k+p)\gamma} \right) \Bigg\} \quad . \end{aligned} \tag{31}$$

#### Recursive Solution for Coefficients Constant in Time

##### For $\mathbf{n}_{\infty,\mathbf{k}}$

We can rewrite $n_{\infty,k}(d_0)$ as a recursive expression, which greatly reduces the effort in calculating $\vec{n}_\infty(d_0)$. At steady state where $\vec{n}_k = \vec{n}_\infty$ (such as when $t \to \infty$), $dn_k/dt \to 0$ and Eq 1 becomes

$$n_{\infty,k}(d_0) = \frac{1}{\alpha(d_0)+k\gamma} \left[\beta_k(d_0) + (k+1)\gamma\, n_{\infty,k+1}(d_0)\right] \quad , \tag{32}$$

which can be calculated recursively starting from $k = M_c$ and then descending to $k = 1$. At $k = M_c$, the value is

$$n_{\infty,M_c}(d_0) = \frac{\beta_{M_c}(d_0)}{\alpha(d_0)+M_c\gamma} \quad , \tag{33}$$

since $n_{\infty,k+1} = 0$ for $k \geq M_c$.

**For $\mathbf{n_k}$**

We have so far been unable to make a recursive form for $\vec{n}$ in Eq (29) where the total number of terms scales as $\mathcal{O}\left(M_c\right)$, but there is a recursive form that reduces the number of terms that must be evaluated to scale as $\mathcal{O}\left(M_c^2\right)$ instead of $\mathcal{O}\left(M_c^3\right)$. We can re-express the innermost sum in Eq (29) in terms of a Gauss hypergeometric function. The Gauss hypergeometric function [3] is

$$ {}_2\mathrm{F}_1(a,b;c;z) = \sum_{p=0}^{\infty} \frac{(a)_p (b)_p \, z^p}{(c)_p \, p!} \quad , \tag{34} $$

and the Pochhammer symbol [3] for integer $p$ is

$$ (x)_p = \begin{cases} x(x+1)\ldots(x+p-1) & \text{if } p>0 \quad , \\ 1 & \text{if } p=0 \quad . \end{cases} \tag{35} $$

Note that ${}_2\mathrm{F}_1(a,b;c;z) = {}_2\mathrm{F}_1(b,a;c;z)$. For the special case that $a=-m$ is a negative integer, like in our case, it is [3]

$$ {}_2\mathrm{F}_1(-m,b;c;z) = \sum_{p=0}^{m} \binom{m}{p} \frac{(-1)^p (b)_p z^p}{(c)_p} \quad . \tag{36} $$

This lets us re-express Eq (29) as

$$ n_k(d_0,t) = n_{\infty,k}(d_0) + z^s \left[U_k(d_0,\vec{\beta}(d_0),z) + V_k(\vec{n}_0(d_0),z)\right] \quad , \tag{37} $$

where

$$ V_k(\vec{y},x) = \sum_{i=k}^{M_c} \binom{i}{k} y_i (1-x)^{i-k} \quad , \tag{38} $$

and

$$ \begin{aligned} U_k(d_0,\vec{y},x) &= -\sum_{i=k}^{M_c} \binom{i}{k} y_i \sum_{p=0}^{i-k} \binom{i-k}{p} \frac{(-1)^p x^p}{\alpha(d_0)+(k+p)\gamma} \\ &= -\frac{1}{\gamma s} \sum_{i=k}^{M_c} \binom{i}{k} y_i \, {}_2\mathrm{F}_1(-(i-k),s;s+1;x) \quad , \end{aligned} \tag{39} $$

and we have defined

$$ s(d_0) = \frac{\alpha(d_0)}{\gamma} + k \quad , \tag{40} $$

$$ z(t) = e^{-(t-t_0)\gamma} \quad . \tag{41} $$

The last definitions mean that $e^{-(\alpha+k\gamma)(t-t_0)} = z^s$. Note that since $t \geq t_0$, $z \in (0,1]$ which means we are in the domain of the Gauss hypergeometric functions of the form in Eq (36)

We need to separate the sum in $U_k(d_0,\vec{y},x)$ into the $i=k$ case and the rest, which is a sum over the same range as for $n_{k+1}$ but with different terms inside. Additionally, $\binom{i}{k} = \frac{k+1}{i-k}\binom{i}{k+1}$ and ${}_2\mathrm{F}_1(0,s;s+1;x) = 1$ Then,

$$U_k(d_0, \vec{y}, x) = -\frac{1}{\gamma s}\left[ y_k + (k+1) \sum_{i=k+1}^{M_c} \binom{i}{k+1} \frac{y_i}{i-k} {}_2\mathrm{F}_1(-(i-k), s; s+1; x) \right] . \quad (42)$$

To build a recursive relationship from $k$ to $k+1$, we need to relate ${}_2\mathrm{F}_1(-(i-k), s; s+1; x)$ to ${}_2\mathrm{F}_1(-(i-k)+1, s+1; s+2; x)$ since $k \to k+1$ changes $s \to s+1$ and $-(i-k) \to -(i-k)+1$. To do this, we will use the contiguous relations/functions which can be used to relate any three Gauss hypergeometric functions of the form ${}_2\mathrm{F}_1(a+m_1, b+m_2; c+m_3; z)$ for different combinations of integers $m_1$, $m_2$, and $m_3$ [3]. We use two from Rakha, Rathie & Chopra (their Eq 1.17 and 1.20) [3] to relate these two hypergeometric functions to each other and ones which can be evaluated exactly. Combining the two contiguous relations,

$$\begin{aligned} {}_2\mathrm{F}_1(-(i-k), s; s+1; x) &= {}_2\mathrm{F}_1(-(i-k)-1, s+1; s+1; x) \\ &\quad + \frac{x}{s+1}\Big[ \\ &\quad (i-k)\,{}_2\mathrm{F}_1(-(i-k)+1, s+1; s+2; x) \\ &\quad + (s+1)\,{}_2\mathrm{F}_1(-(i-k), s+2; s+2; x) \Big] \quad (43) \end{aligned}$$

Now, we have two Gauss hypergeometric functions, with $m \geq 0$, of the form

$$\begin{aligned} {}_2\mathrm{F}_1(-m, c; c; z) &= \sum_{p=0}^{m} \binom{m}{p} \frac{(-1)^p (c)_p z^p}{(c)_p} \\ &= \sum_{p=0}^{m} \binom{m}{p} (-z)^p \\ &= (1-z)^m \quad (44) \end{aligned}$$

Putting this in, we get

$$\begin{aligned} &{}_2\mathrm{F}_1(-(i-k), s; s+1; x) = \\ &\qquad (1-x)^{i-k} + \frac{(i-k)x}{s+1} {}_2\mathrm{F}_1(-(i-k)+1, s+1; s+2; x) \quad . \quad (45) \end{aligned}$$

Inserting this into Eq (42) and stepping the binomial coefficient in the sum with the first term back down to $\binom{i}{k}$,

$$\begin{aligned} U_k(d_0, \vec{y}, x) = -\frac{1}{\gamma s}\Bigg[ & y_k + \sum_{i=k+1}^{M_c} \binom{i}{k} y_i (1-x)^{i-k} \\ & + \frac{(k+1)x}{s+1} \sum_{i=k+1}^{M_c} \binom{i}{k+1} y_i \, {}_2\mathrm{F}_1(-(i-k)+1, s+1; s+2; x) \Bigg] \quad . \quad (46) \end{aligned}$$

We can re-arrange this to be a recursive relationship and combine the first two terms in the brackets into a single sum (the first term is the $i = k$ case of the first sum).

Noting that ${}_2\mathrm{F}_1(0, s; s+1; x) = 1$ for evaluating $U_{M_c}$ with Eq (39), the recursive relationship is

$$U_k(d_0, \vec{y}, x) = \begin{cases} -\frac{y_{M_c}}{\gamma s} & \text{if } k = M_c \quad , \\ \frac{(k+1)x}{s} U_{k+1}(d_0, \vec{y}, x) - \frac{1}{\gamma s} V_k(\vec{y}, x) & \text{otherwise} \quad . \end{cases} \tag{47}$$

The number of terms in evaluating all the $U_k(d_0, \vec{y}, x)$ at a specific $d_0$ and $t$ scales as $\mathcal{O}\left(M_c^2\right)$ with this recursive definition, rather than $\mathcal{O}\left(M_c^3\right)$ if we use the original definition in Eq (39) since the Gauss hypergeometric function itself makes for an inner sum from Eq (36). And therefore we have also made the number of terms required to evaluate $\vec{n}$ at a specific $d_0$ and $t$ scale as $\mathcal{O}\left(M_c^2\right)$, since now there are only single sums in each $n_k$. Additionally, no binomial coefficients are multiplied together anymore which makes it easier to avoid numerical overflow, and the $(-1)^p$ term is gone which causes accuracy problems when doing the inner sum at any given fixed precision.

Looking back at the recursive form of $n_{\infty,k}(d_0)$ in Eq (32) and (33), we can see that

$$n_{\infty,k}(d_0) = -U_k(d_0, \vec{\beta}(d_0), 1) \quad . \tag{48}$$

#### For Time Integral of $\mathbf{n_k}$

Now, we can build a recursive relationship for $\int_{t_0}^{t} n_k dt$ from Eq (30). The double sums can all be re-expressed in terms of $U_k$, getting

$$\begin{aligned} \int_{t_0}^{t} n_k(d_0, v) dv = (t - t_0) n_{\infty,k}(d_0) - U_k(d_0, \vec{n}_0(d_0), 1) \\ + z^s U_k(d_0, \vec{n}_0(d_0), z) + \int_{t_0}^{t} dv\, z(v)^s U_k(d_0, \vec{\beta}(d_0), z(v)) \quad . \end{aligned} \tag{49}$$

Changing integration variables in the last integral from $t$ to $z(v)$, we get

$$\begin{aligned} \int_{t_0}^{t} n_k(d_0, v) dv = (t - t_0) n_{\infty,k}(d_0) - U_k(d_0, \vec{n}_0(d_0), 1) \\ + z^s U_k(d_0, \vec{n}_0(d_0), z) - \frac{1}{\gamma} W_k\left(d_0, \vec{\beta}, z\right) \quad , \end{aligned} \tag{50}$$

where

$$W_k\left(d_0, \vec{y}, x\right) = \int_{1}^{x} dv\, v^{s-1} U_k(d_0, \vec{y}, v) \quad , \tag{51}$$

with a re-use/reassignment of the integration variable.

Then, writing $U_k(d_0, \vec{y}, x)$ out, using Eq (45) to step the Gauss hypergeometric function to $k+1$, and identifying $W_{k+1}\left(d_0, \vec{y}, x\right)$ and $U_k\left(d_0, \vec{y}, x\right)$;

$$
\begin{aligned}
W_k\left(d_0, \vec{y}, x\right) &= -\frac{1}{\gamma s}\int_1^x dv\, v^{s-1}\sum_{i=k}^{M_c}\binom{i}{k} y_i\, {}_2\mathrm{F}_1(-(i-k), s; s+1; v) \\
&= -\frac{1}{\gamma s}\sum_{i=k}^{M_c}\binom{i}{k} y_i\Bigg[\int_1^x v^{s-1}(1-v)^{i-k}dv \\
&\quad + \int_1^x \frac{(i-k)v^s}{s+1}{}_2\mathrm{F}_1(-(i-k)+1, s+1; s+2; v)dv\Bigg] \\
&= -\frac{1}{\gamma s}\sum_{i=k}^{M_c}\binom{i}{k} y_i \sum_{p=0}^{i-k}\binom{i-k}{p}\int_1^x (-1)^p v^{p+s-1}dv \\
&\quad -\frac{k+1}{s}\Bigg[\frac{1}{(s+1)\gamma}\sum_{i=k+1}^{M_c}\binom{i}{k+1} y_i \\
&\quad \bullet \int_1^x v^s{}_2\mathrm{F}_1(-(i-k)+1, s+1; s+2; v)dv\Bigg] \\
&= \frac{k+1}{s}W_{k+1}\left(d_0, \vec{y}, x\right) - \frac{x^s}{\gamma s}\sum_{i=k}^{M_c}\binom{i}{k} y_i\sum_{p=0}^{i-k}\binom{i-k}{p}\frac{(-1)^p x^p}{s+p} \\
&\quad + \frac{1}{\gamma s}\sum_{i=k}^{M_c}\binom{i}{k} y_i \sum_{p=0}^{i-k}\binom{i-k}{p}\frac{(-1)^p}{s+p} \\
&= \frac{1}{s}\left[(k+1)W_{k+1}\left(d_0, \vec{y}, x\right) + x^s U_k\left(d_0, \vec{y}, x\right) - U_k\left(d_0, \vec{y}, 1\right)\right] \quad . \qquad (52)
\end{aligned}
$$

This is recursive since it depends on itself from $k+1$, and $U_k\left(d_0, \vec{y}, x\right)$ and $U_k\left(d_0, \vec{y}, 1\right)$ which are recursive. All that remains is to evaluate it for $k = M_c$, and then Eq (50) is complete. This is

$$
\begin{aligned}
W_{M_c}\left(d_0, \vec{y}, x\right) &= -\frac{1}{\gamma s}\int_1^x dv\, v^{s-1}\sum_{i=M_c}^{M_c}\binom{i}{M_c} y_i\, {}_2\mathrm{F}_1(-(i-M_c), s; s+1; v) \\
&= -\frac{y_{M_c}}{\gamma s}\int_1^x v^{s-1}\, {}_2\mathrm{F}_1(0, s; s+1; v)dv \\
&= -\frac{y_{M_c}}{\gamma s}\int_1^x v^{s-1}\, dv \\
&= \frac{y_{M_c}}{\gamma s^2}\left(1 - x^s\right) \qquad (53)
\end{aligned}
$$
