## Supplementary material for "Risk assessment for airborne disease transmission by poly-pathogen aerosols": S2_appendix

### S2 Appendix. Checking Analytical Solution Against Numerical Solution

The recursive analytical solution for $\vec{n}$ will be compared against a numerical solution. Lets consider, for some fixed $d_0$, the following case.

$$\begin{aligned}
M_c &= 10 \\
\alpha &= \frac{3}{2} \quad [\mathrm{T}]^{-1} \\
\beta &= 50 + 20k \quad [\mathrm{L}]^{-4}\,[\mathrm{T}]^{-1} \\
\gamma &= \frac{1}{10} \quad [\mathrm{T}]^{-1} \\
n_{0,k} &= \left| \frac{M_c}{2} - |4-k| \right|^4 \quad [\mathrm{L}]^{-4}
\end{aligned}$$

where [T] is the unit of time and [L] is the unit of length. This example starts with both some $n_{0,k} < n_{\infty,k}$ and some $n_{0,k} > n_{\infty,k}$ and a source whose strength increases with $k$. The system was solved from $t = t_0 = 0$ [T] to $t = 100$ [T] in steps of $10^{-2}$ [T] for the analytical solution and $10^{-3}$ [T] for the numerical solution. The system of ODEs was solved numerically using the standard Runge-Kutta 4 in IEEE 754 `binary64` floating point (commonly known as `float64` or double precision). The small time step was chosen in order to check that the differences between the two solutions are small. The analytical solution to the concentration densities $n_k(d_0, t)$ over time is shown in the left panel in Fig 1, along with the normalized residual between the analytical and numerical solutions (absolute value of the difference divided by the analytical solution) in the right panel. The concentration densities decay or grow from $\vec{n}_0$ towards $\vec{n}_\infty$ as we expect. The differences between the analytical and numerical calculations are small (less than $10^{-12}$); sometimes reaching the smallest relative differences that can be represented in IEEE 754 `binary64` numbers with their 53 bit mantissas [1], which are $2.2 \times 10^{-16}$ (numerical bigger than analytical by a fraction of $2^{-52}$) and $1.1 \times 10^{-16}$ (numerical smaller than analytical by a fraction $2^{-53}$).

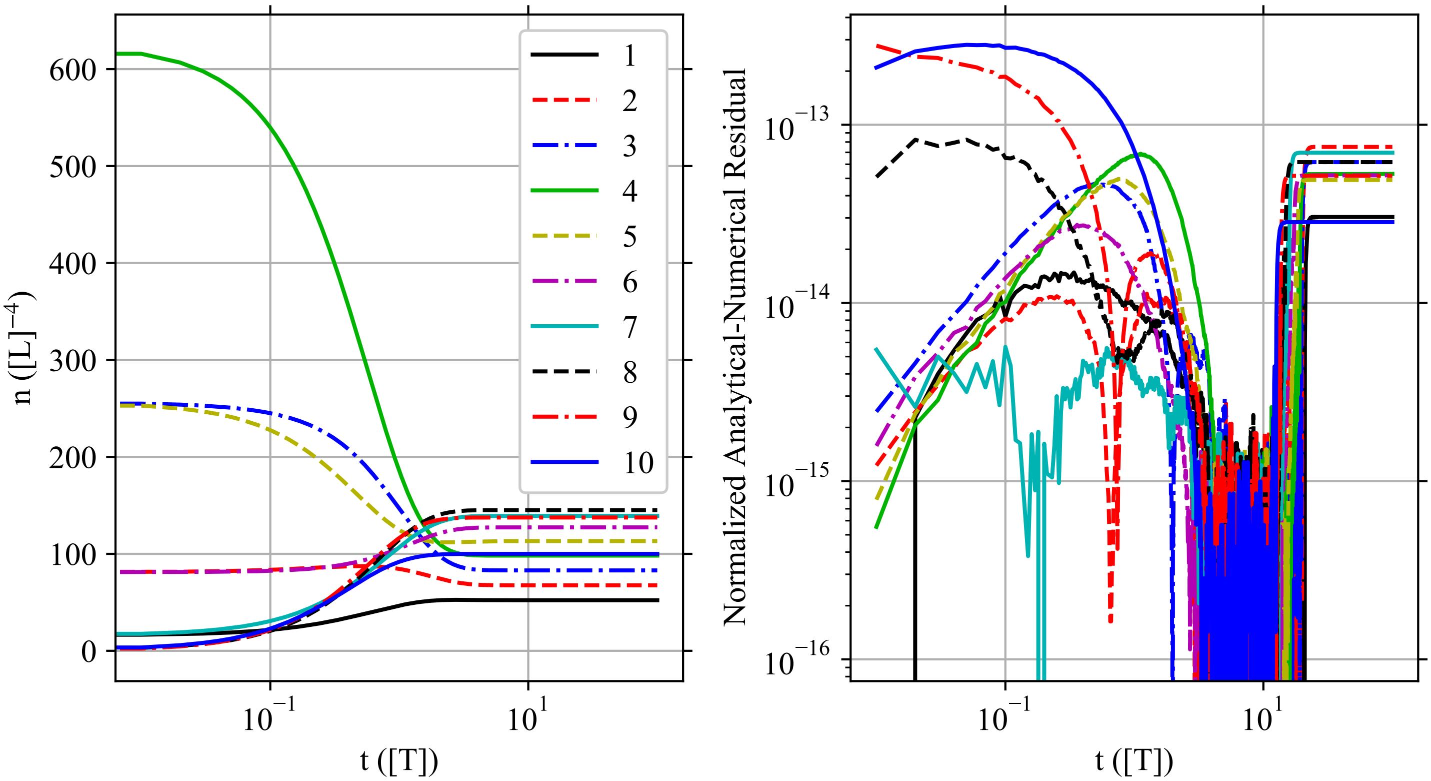


**Fig 1. Numerical Validation of Analytical Solution.** Comparison of the analytical and numerical solutions of $\vec{n}$ for one particular case and $d_0$. (Left) Analytical solution (recursive form) to the infectious aerosol concentration density $n_k(t)$ over time, and (Right) the normalized residual between the analytical and numerical solutions $(|n_{k,analytical} - n_{k,numerical}| / n_{k,analytical}$ over all time steps except $t_0$ where some $n_{0,k}$ are zero. Each $k$ is drawn as a separate line, labeled by the value of $k$. Both panels share the same legend, which is in the Left panel. The numerical solution was done by Runge-Kutta 4 with a time step of $10^{-3}$ [T] using IEEE 754 `binary64` arithmetic.
