## Supplementary material for "Risk assessment for airborne disease transmission by poly-pathogen aerosols": S3_appendix

### S3 Appendix. Binning Diameter

Since each $d_0$ can be treated in isolation of all other $d_0$ in the model's system of equations, we can safely split the $d_0 \in [d_{m,1}, d_M]$ range into finite sized diameter bins and solve each separately and independently of the other bins. We can then apply the general solution to each bin using coefficients averaged over the diameter range in the bin with suitable weights.

Let the $i$'th strictly increasing bin boundary be $d_{b,i}$ for $i \in [0, N_b]$ where $N_b$ is the number of bins, $d_{b,0} = d_{m,1}$, and $d_{b,N_b} = d_M$. The $i$'th bin will refer to the bin $d_0 \in [d_{b,i-1}, d_{b,i})$ for $i \in [1, N_b]$. In the limit that the width of the widest bin goes to zero, we get the exact answer. For finite width bins, the more that $\alpha(d_0, t)$ and $\beta_k(d_0, t)$ vary with respect to $d_0$ in the bin, the less accurate the result from applying binning will be. In practice, this means that we could choose to do a single bin to make it easier to compute the answer but suffer accuracy problems or we could choose a smaller bin width that is small compared to the scale that $\alpha(d_0, t)$ and $\beta_k(d_0, t)$ change over and get good accuracy at the expense of effort. An added benefit of binning is that we can choose different $M_c$ for each bin. In many cases, $M_c$ will be one except for the largest diameter bins.

For any choice of bins, we must determine suitable average values of $n_{0,k}(d_0)$, $n_{r,k}(d_0, t)$, $\alpha(d_0, t)$, and $\beta_k(d_0, t)$. One possible scheme would be

$$\begin{aligned}
\left. n_{0,k} \right|_i &= \left\langle n_{0,k}(d_0) \right\rangle_i \quad , & (1)\\
\left. n_{r,k} \right|_i (t) &= \left\langle n_{r,k}(d_0,t) \right\rangle_i \quad , & (2)\\
\left. \beta_{r,k} \right|_i (t) &= q_r(t) \left\langle n_{r,k}(d_0,t) \right\rangle_i \quad , & (3)\\
\left. \beta_{I,k} \right|_i (t) &= \frac{N_I}{V} \left\langle \lambda_I(t) \left\langle n_{I,k}(d_0,t) \left[1 - E_{I,m,out}(d_0)\right] \right\rangle_i \right\rangle_I \quad , & (4)\\
\left. \beta_k \right|_i (t) &= \left. \beta_{r,k} \right|_i (t) + \left. \beta_{I,k} \right|_i (t) \quad , & (5)\\
\left. \alpha_v \right|_i (t) &= q_v(t) \left\langle E_v \left(w(d_0,t)d_0\right) \right\rangle_i \quad , & (6)\\
\left. \alpha_g \right|_i (t) &= \left\langle \alpha_g \left(w(d_0,t)d_0\right) \right\rangle_i \approx \frac{g\left(\rho_w - \rho_a\right) \left\langle w(d_0,t)^2 \right\rangle_i \left(d_{b,i}^3 - d_{b,i-1}^3\right)}{54 h \rho_a \nu_a \left(d_{b,i} - d_{b,i-1}\right)} \quad , & (7)\\
\left. \alpha_d \right|_i (t) &= \left\langle \alpha_d \left(w(d_0,t)d_0\right) \right\rangle_i \quad , & (8)\\
\left. \alpha_{C,f} \right|_i (t) &= \frac{N_C}{V} \Bigg\langle \lambda_C(t) \Bigg\langle 1 - \left[1 - E_{C,m,in}\left(w(d_0,t)d_0\right)\right] \\
&\quad \bullet \left[1 - E_{C,r}(d_0, w(d_0,t), \lambda_C(t))\right] \left[1 - E_{C,m,out}(d_0)\right] \Bigg\rangle_i \Bigg\rangle_C \quad , & (9)\\
\left. \alpha \right|_i (t) &= \alpha_o(t) + \alpha_r(t) + \left. \alpha_v \right|_i (t) + \left. \alpha_g \right|_i (t) + \left. \alpha_d \right|_i (t) \\
&\quad + \left. \alpha_{I,f} \right|_i (t) + \left. \alpha_{S,f} \right|_i (t) + \left. \alpha_{N,f} \right|_i (t) \quad , & (10)
\end{aligned}$$

where $\alpha_o(t)$ and $\alpha_r(t)$ do not require taking bin averages at all since they don't depend on $d_0$, we have assumed that $w$ is approximately constant with respect to diameter across the bin in evaluating $\left. \alpha_g \right|_i (t)$, taking the average value of $w^2$ over the bin, which is essentially ignoring the effect of surface tension (the quality of the approximation gets better as $d_0$ increases), the notation $\left. F \right|_i (t)$ denotes the bin average value of $F(d_0,t)$ to use for the $i$'th bin, and the bin average of some quantity $\left\langle F(d_0,t) \right\rangle_i$ is taken by doing

$$\left\langle F(d_0,t) \right\rangle_i \equiv \frac{1}{d_{b,i} - d_{b,i-1}} \int_{d_{b,i-1}}^{d_{b,i}} F(\phi,t) d\phi \quad . \qquad (11)$$

If a bin is small enough compared to the variation in $F(d_0,t)$ with respect to $d_0$, one could just approximate the average as the value of $F$ for an arbitrary $d_0 \in [d_{b,i-1}, d_{b,i}]$.

To get the actual number of aerosols $\mathcal{N}_{a,i}$ in each bin, we need to multiply by the bin width, which is

$$\mathcal{N}_{a,i}(t) = \left(d_{b,i} - d_{b,i-1}\right) \left. n_k \right|_i (t) \quad . \qquad (12)$$

We also need to get the average **aerosol** dose in each bin by suitably averaging $\mu_{j,k}(t)$ over the bin. One possible scheme is

$$\left. \mu_{j,k} \right|_i (t) = \int_{t_0}^{t} dv \, \left\langle E_{S,r,j}(d_0) \left[1 - E_{S,m,in,j}\left(w(d_0,v)d_0\right)\right] \right\rangle_i \lambda_{S,j}(v) \mathcal{N}_{a,i}(v) \quad , \qquad (13)$$

where $\mathcal{N}_{a,i}(t)$ has taken the place of $\left. n_k \right|_i (t)$

Regardless of the scheme, the average dose for each multiplicity is calculated by summing over the bins as

$$\mu_{j,k}(t) = \sum_{i=1}^{N_b} \left. \mu_{j,k} \right|_i (t) \quad . \qquad (14)$$
