## Supplementary material for "Risk assessment for airborne disease transmission by poly-pathogen aerosols": S4_appendix

### S4 Appendix. $\mathbf{M_c}$ Heuristic for Infectious People Derivation

The $M_c$ heuristic for production by all infectious people is

$$\mathcal{H}_{I,k}(t) \equiv k \int_{d_-}^{d_+} d\phi \, A_S(\phi, t) \sum_{j=1}^{N_I} \lambda_{i,j}(t) n_{I,j,k}(\phi, t) \left[1 - E_{I,m,out,j}(\phi)\right] \quad . \tag{1}$$

For this heuristic, we can simplify $J_{I,M_c}(t)$ since $n_{I,j,k}(d_0, t)$ is the product of a scalar and a Poisson probability until $d_{m,k} > d_0$ after which it is zero. Now $n_{I,j,k}(d_0, t) = 0$ for $k > K_m(d_0)$. The sums and the integrals all commute, so we can change their order. We then get inner sums of the following form in both the numerator and the denominator

$$\begin{aligned} Z(\mu, m, K_m) &= \sum_{k=m}^{K_m} k P_P(\mu, k) \\ &= \sum_{k=m}^{K_m} k e^{-\mu} \frac{\mu^k}{k!} \\ &= \mu \sum_{k=m}^{K_m} e^{-\mu} \frac{\mu^{k-1}}{(k-1)!} \\ &= \begin{cases} 0 & \text{if } m > K_m \\ \mu \left[C_P(\mu, K_m - 1) - C_P(\mu, m - 2)\right] & \text{otherwise} \end{cases} \\ &= \mu \left[C_P(\mu, K_m - 1) - C_P(\mu, \min(m - 2, K_m - 1))\right] \quad , \end{aligned} \tag{2}$$

where $m$ is the starting index and $C_P(\mu, k)$ is the CDF (Cumulative Distribution Function) of the Poisson distribution with mean $\mu$ for count $k$, with $C_P(\mu, k) = 0$ for $k < 0$. Note that

$$Z(\mu, 1, K_m) = \mu C_P(\mu, K_m - 1) \quad . \tag{3}$$

Then

$$
\begin{aligned}
J_{I,M_c}(t) = \Bigg\{ \int_{d_-}^{d_+} & d\phi \, A_S(\phi,t) \\
& \bullet \sum_{j=1}^{N_I} \lambda_{I,j}(t) \left[1 - E_{I,m,out,j}(\phi)\right] \rho_j(\phi,t) Z \left( \langle k \rangle (\phi,t)_j \, , M_c + 1, K_m \right) \Bigg\} \\
& \qquad \Bigg/ \Bigg\{ \int_{d_-}^{d_+} d\phi \, A_S(\phi,t) \\
& \qquad \sum_{j=1}^{N_I} \lambda_{I,j}(t) \left[1 - E_{I,m,out,j}(\phi)\right] \rho_j(\phi,t) Z \left( \langle k \rangle (\phi,t)_j \, , 1, K_m \right) \Bigg\} \quad . \qquad (4)
\end{aligned}
$$

Our heuristic is strictly for all of the $J_{h,M_c}$. But, we could widen the set of heuristics by doing one for each infectious person. If we also replace the integral by an evaluation at $d_+$, the largest $d_0$ in the interval, and ignore the filtering efficiency of the infectious person's mask on the way out; then there exists some $j \in I$ for which $M_{c,I,j} \geq M_{c,I}$, meaning that the maximum $M_{c,I,j}$ is an overestimate for $M_{c,I}$. If we use this heuristic, we could end up having to use a larger $M_c$ than we really need; but this method has the advantage of easy calculation. This new heuristic for the $j$'th infected person is

$$
\begin{aligned}
\mathcal{J}_{I,M_{c,I,j},j}(t) &= \frac{Z \left( \langle k \rangle (d_+,t)_j \, , M_{c,I,j} + 1, K_m(d_+) \right)}{Z \left( \langle k \rangle (d_+,t)_j \, , 1, K_m(d_+) \right)} \\
&= 1 - \frac{C_P \left( \langle k \rangle (d_+,t)_j \, , \min(M_{c,I,j}, K_m(d_+)) - 1 \right)}{C_P \left( \langle k \rangle (d_+,t)_j \, , K_m(d_+) - 1 \right)} \quad , \qquad (5)
\end{aligned}
$$

where $K_m(d_0)$ is evaluated for $d_0 = d_+$.

With the threshold $T \in (0,1]$, we get

$$
\begin{aligned}
& C_P \left( \langle k \rangle (d_0,t)_j \, , \min(M_{c,I,j}, K_m(d_+)) - 1 \right) \\
& \qquad \geq (1-T) \, C_P \left( \langle k \rangle (d_0,t)_j \, , K_m(d_+) - 1 \right) \quad , \qquad (6)
\end{aligned}
$$

and then must find the smallest $M_{c,I,j} \leq K_m(d_+)$ for which this is true. This value exists because $M_{c,I,j} = K_m(d_+)$ trivially makes the statement true. Then,

$$
M_{c,I,j}(d_+,T) = 1 + C_P^{-1} \left( \langle k \rangle (d_+,t)_j \, , (1-T) \, C_P \left( \langle k \rangle (d_+,t)_j \, , K_m(d_+) - 1 \right) \right) \quad , \qquad (7)
$$

where $C_P^{-1}(\mu, c)$ is the inverse CDF to find the smallest $k$ for which $C_P(\mu, k) \geq c$.
