## Supplementary material for "Risk assessment for airborne disease transmission by poly-pathogen aerosols": S5_appendix

### S5 Appendix. Numerical Considerations

If $\alpha$, $\beta_k$, $\gamma$, and $w$ are all constant with respect to time; the model has both an explicit and recursive solution for the concentration density and dose. Otherwise, it may not be possible to get a closed form analytical solution from the general solution and one would need to solve the model numerically. Of course, even with an analytical solution, one can solve it numerically. There are a number of numerical pitfalls with both the analytical and numerical solutions, which arise as $M_c$ becomes large.

For a numerical solution, the time step of integration must be small compared to the smallest time scale in the model. The smallest time scale could be the time scale on which $\alpha$, $\beta_k$, $\gamma$, and/or $w$ change over time; but for large enough $M_c$ it will always be time scale of the total sink for $k = M_c$ aerosols. The total sink for $n_k$ is $-\alpha(d_0, t) - k\gamma(t)$, which scales linearly in $M_c$ for $n_{M_c}$ when $M_c$ is large enough. The time step $\delta t$ must be $\delta t < (\alpha + M_c\gamma)^{-1}$. Thus the total number of timesteps $N_t$ scales as $N_t \sim M_c$ for large $M_c$. Since the total number of terms scales linearly in $M_c$ as long as $\partial n_k / \partial t$ is not calculated in vector-matrix form or $\mathbf{A}$ is stored in a sparse format, the total computational effort scales as $\mathcal{O}\left(M_c^2\right)$. Also note, the model is stiff when $M_c\gamma(t) \gg \alpha(d_0, t) + \gamma(t)$ since the total sink timescale for $k = M_c$ is much shorter than the total sink timescale for $k = 1$.

For the analytical solution for coefficients constant in time; there are different difficulties for the explicit solution and the recursive solution. The number of terms scales as $\mathcal{O}\left(M_c^3\right)$ in the explicit solution and $\mathcal{O}\left(M_c^2\right)$ in the recursive solution, since there are $M_c$ equations and they have double and single sums respectively, with each sum scaling as $M_c$.

The analytical solutions have additional difficulties — avoiding numerical overflow and maintaining accuracy. The problem is the binomial coefficients $\binom{i}{k}$ and $\binom{i-k}{p}$ where $i \to M_c$ and $p \in [0, i - k]$. They can be calculated naively by computing each factorial and then doing the multiplication and division ($k!$ will be the largest); or carefully with cancellation handled explicitly or using logarithms of the gamma function, which overflows later. And then even if all the binomial coefficients don't overflow, their products and sums can still overflow (the explicit version is worse than the recursive version in this regard due to the products of binomial coefficients).

To see this, we will find the upper bound for $V_k\left(\vec{y}, x\right)$ in the recursive solution. Consider its formula

$$V_k\left(\vec{y},x\right) = \sum_{i=k}^{M_c} \binom{i}{k} y_i (1-x)^{i-k} \quad . \tag{1}$$

In the model, all the elements of $\vec{y}$, which is always either $\vec{\beta}$ or $\vec{n}_0$, are non-negative. Since $x \in [0,1]$, $1-x \in [0,1]$ and is therefore also non-negative. This means that all elements in the sum in $V_k$ are non-negative, meaning $V_k \geq 0$ and thus we only need to find the upper bound to determine the risk of overflow (the lower bound, zero, is not a worry). Now, $y_i \leq \max\left(\vec{y}\right)$ for all $i$ where $\max\left(\vec{y}\right)$ shall denote the maximum element of $\vec{y}$. Since $x \in [0,1]$, $0 \leq (1-x)^{i-k} \leq 1$. Then we just need to get an upper bound for the binomial coefficients. The binomial coefficient $\binom{j}{m} = k!/m!(j-m)!$ is at its greatest value for fixed $j$ when $m = \lfloor j/2 \rfloor$ or $m = \lceil j/2 \rceil$ where $\lfloor \cdot \rfloor$ and $\lceil \cdot \rceil$ denote the floor and ceil operators respectively. This value is

$$\binom{j}{\lfloor j/2 \rfloor} = \binom{j}{\lceil j/2 \rceil} = \frac{j!}{\lfloor j/2 \rfloor ! \lceil j/2 \rceil !} \quad . \tag{2}$$

This will be maximized when $i = j = M_c$. Then, the upper bound for $V_k$ is

$$V_k\left(\vec{y},x\right) \leq \sum_{i=k}^{M_c} \frac{M_c!}{\lfloor M_c/2 \rfloor ! \lceil M_c/2 \rceil !} \max\left(\vec{y}\right) = \frac{(M_c+1-k) M_c! \max\left(\vec{y}\right)}{\lfloor M_c/2 \rfloor ! \lceil M_c/2 \rceil !} \quad . \tag{3}$$

For any $M_c$, the largest upper bound for any of the $V_k$ is found for $k=1$, so the upper bound we need to worry about is

$$V_k\left(\vec{y},x\right) \leq \frac{M_c M_c! \max\left(\vec{y}\right)}{\lfloor M_c/2 \rfloor ! \lceil M_c/2 \rceil !} \quad . \tag{4}$$

Now, the factorial function has the bounds [1]

$$\sqrt{2\pi} m^{m+\frac{1}{2}} e^{-m} \exp\left[\frac{1}{12m+1}\right] < m! < \sqrt{2\pi} m^{m+\frac{1}{2}} e^{-m} \exp\left[\frac{1}{12m}\right] \quad . \tag{5}$$

We will still have an upper bound for $V_k$ if we use the factorial lower bounds in place of the factorials in the denominator of Eq (4) and the factorial upper bound for the factorial in the numerator. This leads to

$$V_k\left(\vec{y},x\right) < \frac{\max\left(\vec{y}\right) M_c^{M_c+\frac{3}{2}} \exp\left[\frac{1}{12M_c} + \left\lfloor \frac{M_c}{2} \right\rfloor + \left\lceil \frac{M_c}{2} \right\rceil\right]}{\sqrt{2\pi} \left\lfloor \frac{M_c}{2} \right\rfloor^{\left\lfloor \frac{M_c}{2} \right\rfloor + \frac{1}{2}} \left\lceil \frac{M_c}{2} \right\rceil^{\left\lceil \frac{M_c}{2} \right\rceil + \frac{1}{2}} \exp\left[M_c + \frac{1}{12\lfloor \frac{M_c}{2} \rfloor + 1} + \frac{1}{12\lceil \frac{M_c}{2} \rceil + 1}\right]} \quad . \tag{6}$$

If we take the $\log_2$, we get the value of the base-2 exponent. Taking the $\log_2$ of both sides,

$$\begin{aligned} \log_2 V_k\left(\vec{y},x\right) < {} & \log_2\left[\max\left(\vec{y}\right)\right] + \left(M_c + \frac{3}{2}\right) \log_2\left(M_c\right) \\ & - \left(\left\lfloor \frac{M_c}{2} \right\rfloor + \frac{1}{2}\right) \log_2 \left\lfloor \frac{M_c}{2} \right\rfloor - \left(\left\lceil \frac{M_c}{2} \right\rceil + \frac{1}{2}\right) \log_2 \left\lceil \frac{M_c}{2} \right\rceil \\ & + \left(\log_2 e\right) \left(\frac{1}{12M_c} - \frac{1}{12\lfloor \frac{M_c}{2} \rfloor + 1} - \frac{1}{12\lceil \frac{M_c}{2} \rceil + 1}\right) \\ & - \frac{1}{2} - \frac{1}{2} \log_2 \pi \quad , \end{aligned} \tag{7}$$

where we have used the fact that $\lfloor M_c/2 \rfloor + \lceil M_c/2 \rceil = M_c$. To represent all the $V_k$ for a particular $M_c$ without overflowing, a floating point format's maximum supported base-2 exponent, $emax$ (using the same notation as the IEEE 754 standard [2]), must be at least this value ($emax \geq \log_2(V_k)$). But, even if the value $\log_2(V_k)$ might not overflow with this minimum value, calculating the binomial coefficient when $\max(\vec{y}) < 1$ could overflow before the multiplication drops the magnitude. So for the minimum $emax$, we must replace the $\max(\vec{y})$ with $\max[1, \max(\vec{y})]$ and we get

$$emax(\vec{y}) \geq \Bigg\lceil \log_2[\max[1, \max(\vec{y})]] + \left(M_c + \frac{3}{2}\right) \log_2(M_c) - \left(\left\lfloor \frac{M_c}{2} \right\rfloor + \frac{1}{2}\right) \log_2 \left\lfloor \frac{M_c}{2} \right\rfloor - \left(\left\lceil \frac{M_c}{2} \right\rceil + \frac{1}{2}\right) \log_2 \left\lceil \frac{M_c}{2} \right\rceil + (\log_2 e)\left(\frac{1}{12 M_c} - \frac{1}{12\lfloor \frac{M_c}{2} \rfloor + 1} - \frac{1}{12\lceil \frac{M_c}{2} \rceil + 1}\right) - \frac{1}{2} - \frac{1}{2}\log_2 \pi \Bigg\rceil \quad . \tag{8}$$

To see how much of an overestimate this is for $emax$, let's compare it to $\log_2 \max(V_k)$ for a few simple cases. First, lets compare it to $V_k\left(\vec{1}, 0\right)$ since $\vec{y} = \vec{1}$ has all elements equal to the maximum element, $x = 0$ maximizes $(1-x)^{i-k}$, and all terms in the sum are integers. In Fig 1 (left panel), $\log_2 \max(V_k)$ is compared to $emax$ from Eq (8) and the $emax$ values for the four smallest IEEE-754-219 binary floating point formats [2]. We calculated $\max(V_k)$ using variable sized integers before being converting to multi-precision floating point numbers for taking the $\log_2$ with the largest supported exponent with the GNU Multiple Precision Floating-point Reliable Library (MPFR, see `https://www.mpfr.org`) and the GNU Multiple Precision Arithmetic Library (GMP, see `https://gmplib.org`) in Python using the gmpy2 package (`https://pypi.org/project/gmpy2`). The minimum $emax$ from Eq (8) is only barely larger in a logarithmic sense than $\log_2 \max(V_k)$ except for small $M_c$, so it isn't an excessive overestimate of the required $emax$.

Second, we will compare it to typical $\vec{y}$ used in the model. We will base $\vec{y}$ on $\vec{\beta}_{I,k}$ for single infectious person but remove all of the environment parameters and person specific parameters except for the expected average multiplicity $\langle k \rangle_j = \frac{\pi}{6} d_0^3 \rho_{p,j}$. We shall use the vector

$$\psi_k = \frac{V}{\rho_j \lambda_{I,j} S_{I,m,out,j}} \beta_{I,k} = P_P\left(\langle k \rangle_j, k\right) \quad . \tag{9}$$

For a range of $\langle k \rangle_j$, $M_c$ was determined with the single infectious person production heuristic $M_{c,I,j}$ for thresholds $T$ of $10^{-1}$ and $10^{-9}$. Then the $V_k\left(\vec{\psi}, 0\right)$ were calculated in binary floating point with a 256 bit mantissa, $emax = 32768$, and minimum exponent $emin = -32767$ with gmpy2, MPFR, and GMP as before. The binomial coefficients were calculated exactly before conversion to floating point. To improve the accuracy, the sum in Eq (1) was done as a sorting sum where all the terms were sorted in a list, the two smallest terms removed, those terms added together and inserted back into list; and this process repeated till only a single term (the total sum) remained. The right panel of Fig 1 compares $\log_2 \max(V_k)$ against the $emax$ calculated from Eq (8) and the maximum exponents of some of the smallest IEEE 754 binary floating point formats. We can see that Eq (8) overestimates the required $emax$ by quite a bit in a logarithmic

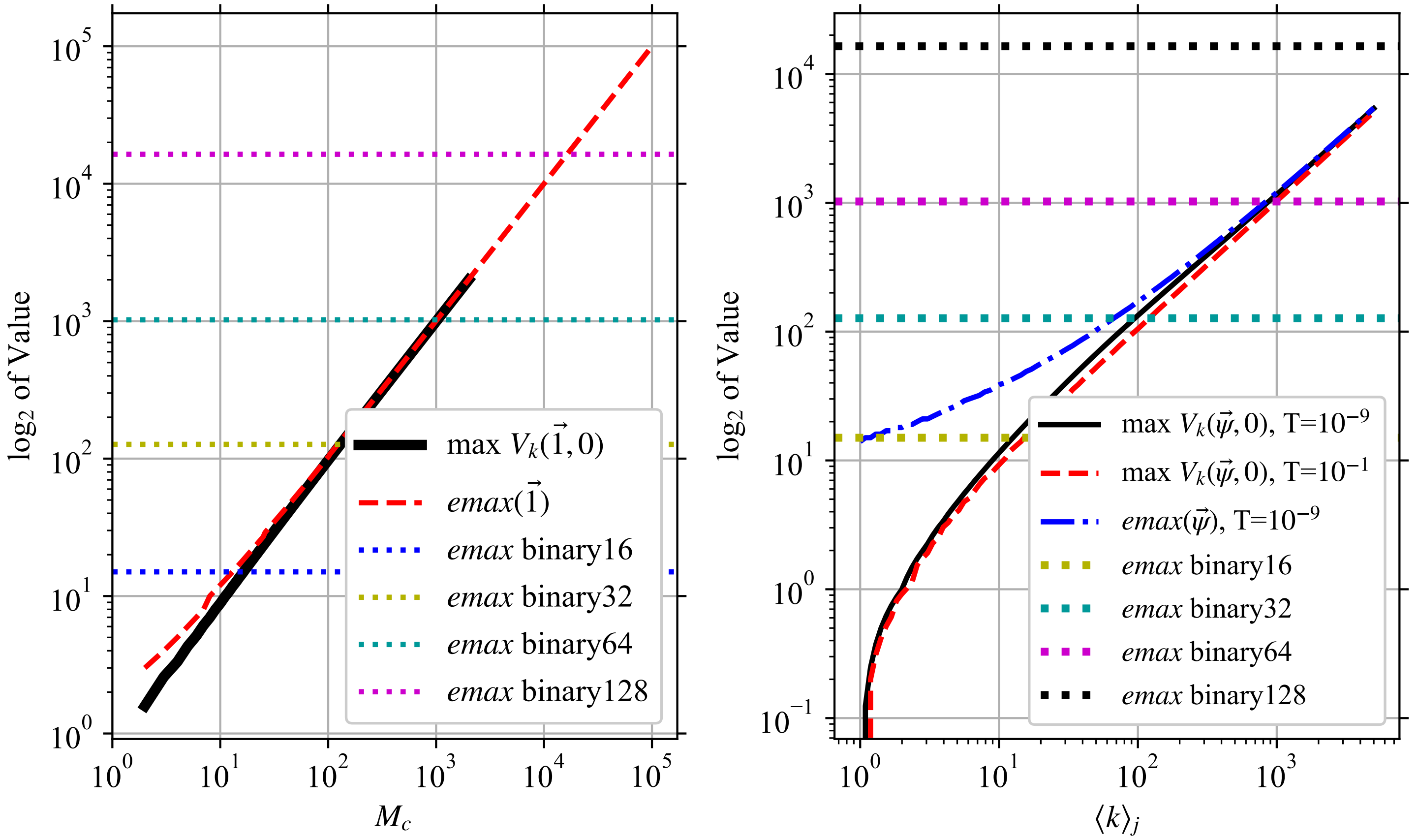


**Fig 1. Comparison of Maximum $V_k$ Values to Required emax Overestimate And Floating Point Format Limits.** $\log_2 \max(V_k)$ is compared to the overestimate of the required $emax(\vec{y})$ from Eq (8) as well as the maximum exponents supported by the four smallest IEEE-754-2019 binary floating point formats [2]. Note that the x87 FPU 80-bit floating point format has the same maximum as `binary128` [3]. (Left) For $\vec{y} = \vec{1}$ and $x = 0$ as a function of $M_c$. (Right) For $\vec{y} = \vec{\psi}$ from Eq (9) and $x = 0$ as a function of the average multiplicity $\langle k \rangle_j = \frac{\pi}{6} d_0^3 \rho_{p,j}$.

sense for $\langle k \rangle_j < 10$, but not by much in a logarithmic sense for larger $\langle k \rangle_j$. For small $\langle k \rangle_j$, it suggests an overkill $emax$ but `binary16` (half precision) and `binary32` (single precision) are typically the smallest floating point formats with hardware support on most computers and the bulk of the computational effort is spent on diameter bins with the largest $M_c$, so there isn't much reason to use a smaller format for small $\langle k \rangle_j$. `binary64` (double precision) is sufficient for $\vec{\psi}$ in this case up to $\langle k \rangle_j = 1000$. The exact values of other terms in $\vec{\beta}_I$ would determine if `binary64` would be safe for that expected multiplicity with $\vec{\beta}_I$. The extra $emax$ requirement would additively increase by $\log_2 \left[\max\left(1, \frac{1}{V} \rho_j \lambda_{I,j} S_{I,m,out,j}\right)\right]$. `binary128` and x87 FPU 80-bit floating point numbers which have the same $emax$ [3] would have some headroom for the magnitude of the other coefficients in $\vec{\beta}_I$ even for $\langle k \rangle_j = 10^4$.

To investigate the required floating point precision to calculate $n_k$, we will look at the required precision in bits required to calculate all $U_k(d_0, \vec{y}, x)$ for a given $M_c$ to within a specified relative tolerance $\delta$ of the exact values. We chose

$$
\begin{aligned}
\alpha &= 1 \quad , \\
\gamma &= 3 \quad , \\
\vec{y} &= \vec{1} \quad , \\
x &\in \left\{0, \frac{5}{13}, 1\right\} \quad ,
\end{aligned}
$$

which makes $U_k\left(d_0, \vec{y}, x\right)$ a rational number for any $M_c$. For several $M_c$, the $U_k\left(d_0, \vec{y}, x\right)$ were calculated exactly using variable sized rational arithmetic. Then, the smallest floating point precision was found for which the maximum relative error in the $U_k\left(d_0, \vec{y}, x\right)$ calculated in floating point in that precision (note, the binomial coefficents were calculated first as variable sized integers and then converted to floating point) is less than $\delta$ using the explicit and recursive formulas for $U_k\left(d_0, \vec{y}, x\right)$. The calculations were done using gmpy2, MPFR, and GMP as before. The required precisions for $\delta$ of 10%, 10 PPM (Parts per Million), and 1 PPB (Parts per Billion) are shown in Fig 2 and compared to the precisions of various standard floating point formats.

Using the explicit formula, even quadruple precision (`binary128`) is insufficient by $Mc = 80$ for this $\vec{y}$ even for a tolerance of 10%. But using the recursive formula, double precision (`binary64`) is good enough for a tolerance of 1 PPB even for the largest $M_c = 5000$ that was checked. From this and the recursive solution's number of terms scaling as $\mathcal{O}\left(M_c^2\right)$ instead of $\mathcal{O}\left(M_c^3\right)$, the recursive solution is much more amenable to calculations than the explicit solution.

Considering both Fig 1 and 2, double precision (`binary64`) seems to be suitable, depending on the largest values in the $\vec{y}$, for moderate $M_c$ of a few hundred. To go to $M_c$ of a few thousand, quadruple precision (`binary128`) or x87 FPU 80-bit floating point numbers are required. Above this, either multi-precision floating point with higher exponents must be used or the model must be solved numerically.

The number of terms to compute scales quadratically in $M_c$ for both the numerical solution and recursive analytical solution, and cubically for the explicit analytical solution; and the analytical solutions have the additional problems of avoiding numerical overflow as well as the computational effort to calculate some terms increasing with $M_c$. For any particular fixed size number format, there is an $M_c$ above which the analytical result will overflow and one must solve the model numerically instead.

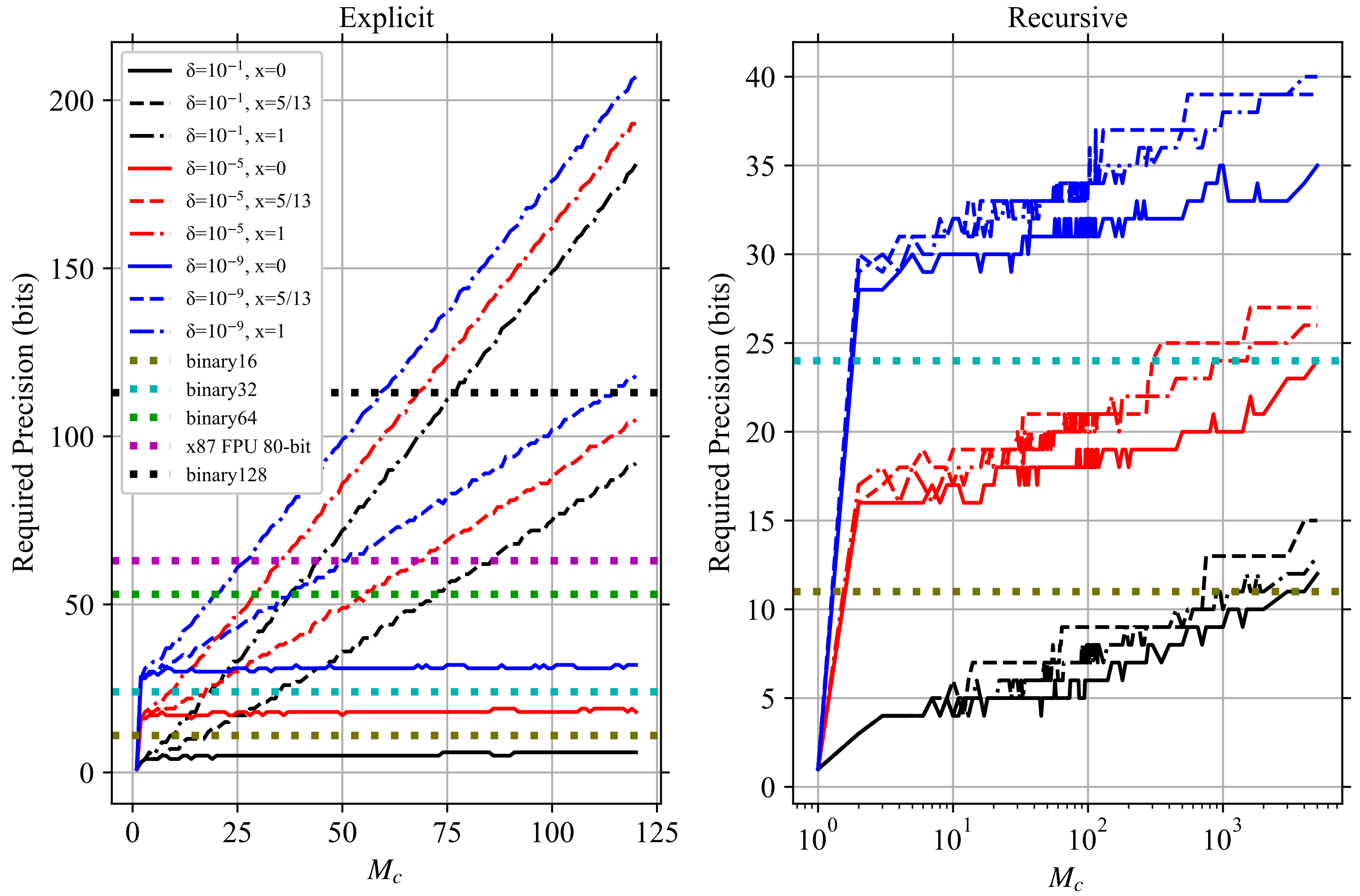


**Fig 2. Required Floating Point Precision to Calculate $\mathbf{U_k}$.** The required floating point precision in bits to get the largest relative error in the $U_k(d_0, \vec{y}, x)$ to be less than the tolerance $\delta$ for three values of $x$ using the (Left) explicit formula and (Right) recursive formula. Horizontal lines show the precisions provided by the four smallest IEEE-754-2019 binary floating point formats [2] and the x87 FPU 80-bit floating point format [3]. Both panels share the same legend, which is in the left panel.
