## Supplementary figures and images for "Risk assessment for airborne disease transmission by poly-pathogen aerosols"

### S6_Fig

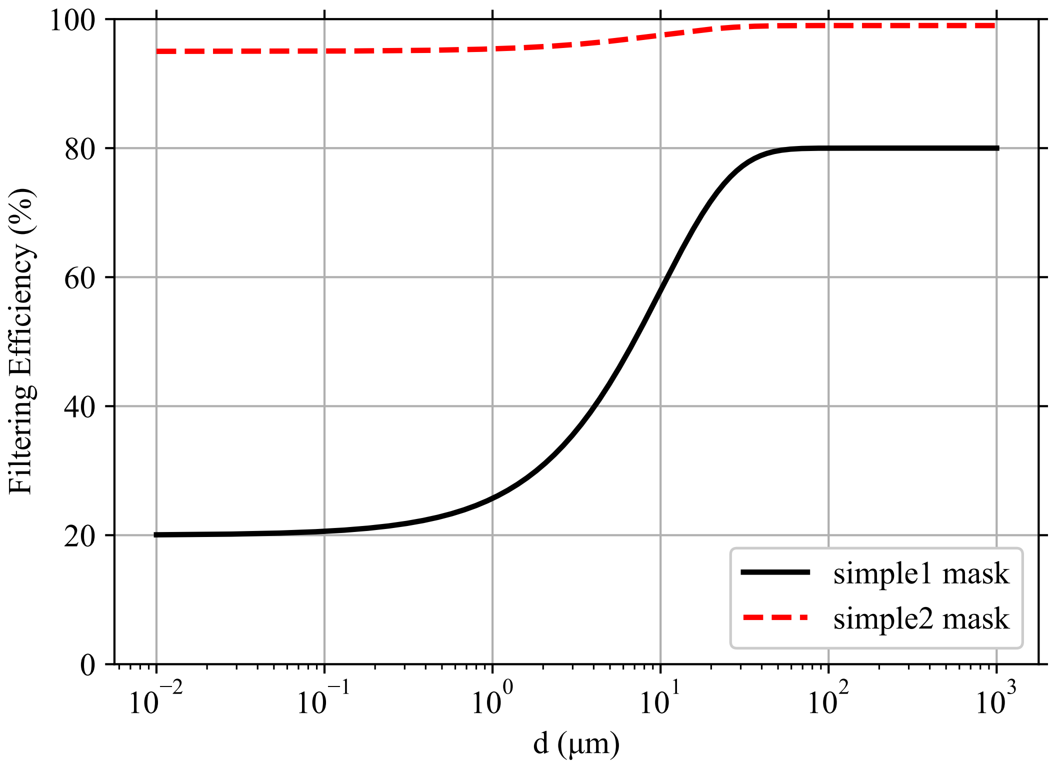

### S7_appendix

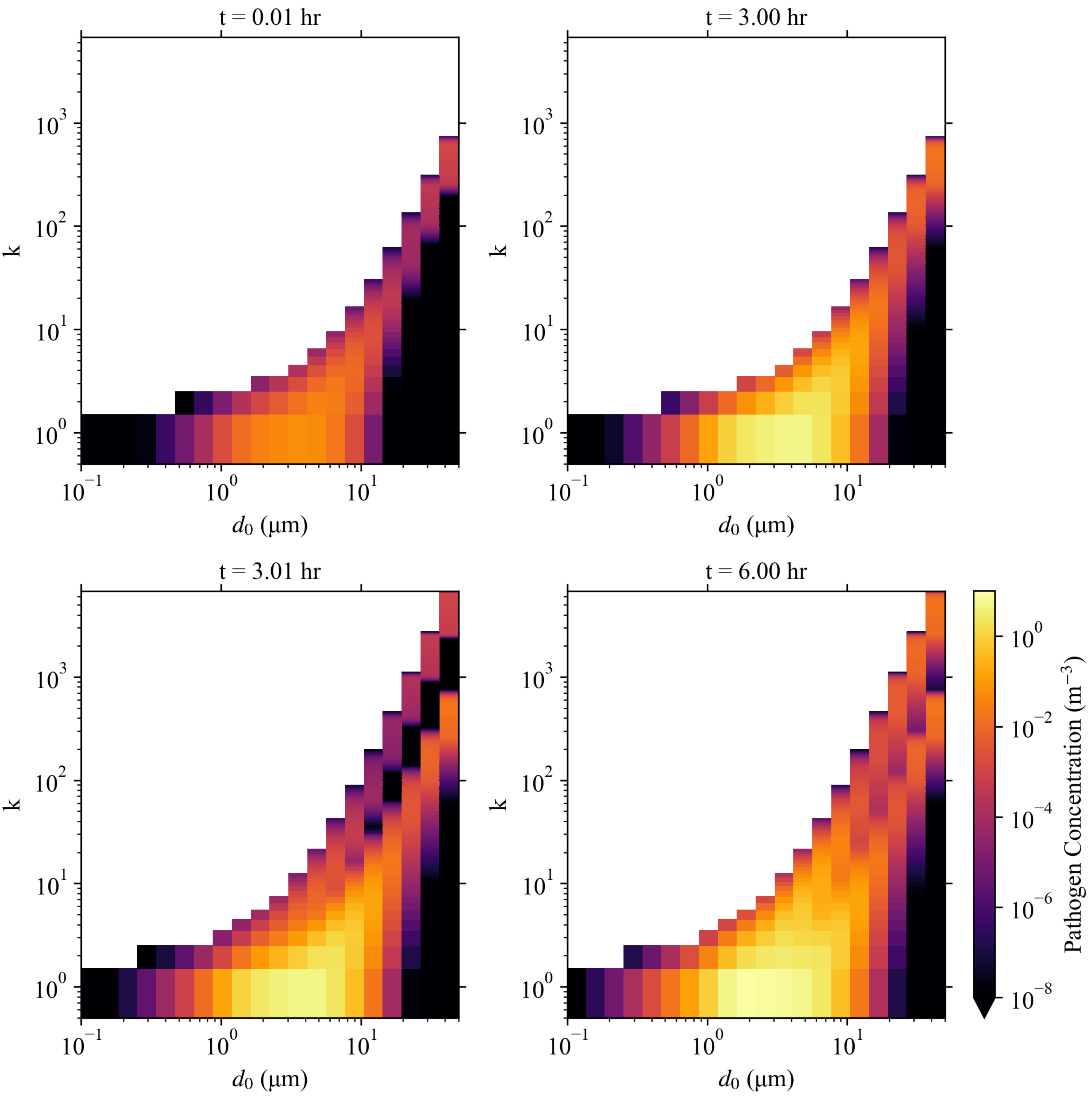

### S8_Fig

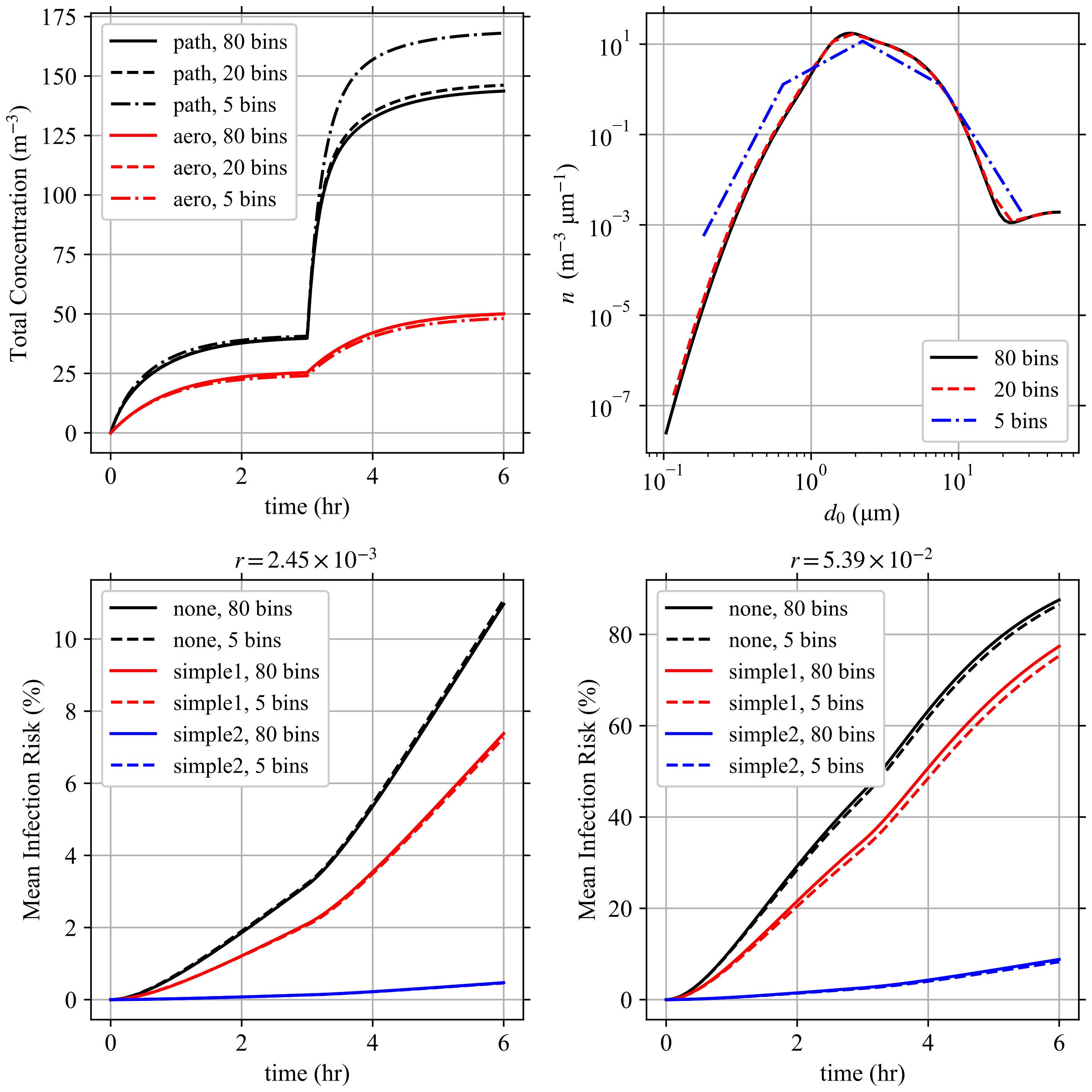
